# Remote Physiologic Monitoring and Principal Care Management for Chronic Retinal Diseases: Results from over 80,000 Encounters

**DOI:** 10.64898/2026.02.27.26347265

**Authors:** Sonia Dhoot, David Boyer, Robert Avery, Glenn Stoller, Stephen Couvillion, Philip Ferrone, Patrick Crane, Tsontcho Ianchulev, Earnest P Chen

**Affiliations:** Harvard Eye Associates, California, USA; Retina Vitreous Associates Medical Group, Beverly Hills, CA, USA.; Department of Ophthalmology, Perelman School of Medicine, University of Pennsylvania, Philadelphia, PA; Ophthalmic Consultants of Long Island, Long Island, NY, USA.; California Retina Consultants, Bakersfield, California; Department of Ophthalmology, Northwell Health Eye Institute and Vitreoretinal Consultants of New York, Great Neck, New York.; Department of Ophthalmology, Icahn School of Medicine at Mount Sinai, New York, USA; New York Eye and Ear Infirmary at Mount Sinai, New York, USA

**Keywords:** Telemedicine, Remote Monitoring, Ophthalmology, Retina, Macustat

## Abstract

**Purpose:** Timely detection of disease activity in chronic retinal diseases improves visual outcomes but is limited by the lack of validated systems for continuous monitoring and care management. We evaluated the real-world performance of an integrated remote physiologic monitoring and principal care management program (RemoniHealth®) using a self-administered multimodal retinal function test (Macustat®) for home monitoring.

**Methods:** This single-arm real-world intervention study was conducted across 33 retina practices. A total of 2,216 adults with chronic retinal diseases performed weekly home retinal function testing with integrated care management support. Primary endpoints included the annualized rate of disease progression detection, time to intervention after first flag, true positive rate, and patient adherence. Descriptive statistics and data analyses were analyzed using chi-square tests and Clopper–Pearson confidence intervals.

**Results:** Participants contributed 82,644 encounters and 16,805 patient-months of monitoring. The program generated 241 alerts, including 101 Macustat flags and 135 care management prompts. Among 73 adjudicated flags, 56 were true positives and 17 false positives (PPV 76.7%). The annualized detection rate was 4 per 100 patient-years. Of confirmed events, 93% led to intravitreal injection or other major management change. Mean adherence was 72.1%, and patients with ≥80% adherence had higher odds of true positivity.

**Discussion:** This RPM–PCM model achieved high engagement and meaningful detection of asymptomatic progression between visits, supporting the value of home monitoring for timely intervention.

**Translational Relevance:** These findings support scalable integration of home vision testing and care management into routine retinal practice to enable earlier intervention and improved continuity of care.

## Introduction

The management of chronic retinal diseases, such as age-related macular degeneration (AMD), diabetic retinopathy (DR), and retinal vein occlusion (RVO), represents a significant challenge due to their progressive nature, treatment burden, and potential for severe vision loss if not monitored and treated promptly.^1–4^ These conditions affect millions globally, with AMD alone estimated to impact over 196 million people worldwide by 2020, a figure projected to rise to 288 million by 2040 due to aging populations.^8^ The advent of anti-vascular endothelial growth factor (anti-VEGF) therapies has revolutionized treatment outcomes, yet their efficacy is highly dependent on timely detection and intervention, often requiring frequent monitoring that exceeds the capacity of traditional clinic-based care models.^6–9,19^

To address the gap between the need for more frequent monitoring and the limitations of episodic in-office visits, remote physiologic monitoring (RPM) and principal care management (PCM) have emerged as complementary approaches, leveraging digital health technologies to provide care continuity, patient engagement and disease monitoring for timely progression detection and treatment. Such programs are becoming more common in many areas of medicine from specialty care to primary care where both RPM and PCM are showing clinical utility and population health benefits.^29–34^ For example, several recent syntheses and trials show RPM can cut heart failure hospitalizations and improve management, especially when devices feed data to a clinical team and trigger action.^31^ Meta-analyses through of studies between 2024 and 2025 show small-to-moderate HbA1c reductions with remote/connected glucose monitoring and app-enabled follow-up (≈0.3–0.4% absolute A1c drop).^30^ Within maternal health, remote postpartum BP monitoring programs (at a safety-net hospital) reliably captured hypertension and supported follow-up.^33^ And for gestational diabetes, telemedicine/RPM meta-analyses suggest improved glycemic control vs paper-based/standard care in several trials.^34^

The integration of RPM into retinal disease management has been propelled by advancements in telemedicine, spurred notably by the COVID-19 pandemic, which highlighted the limitations of in-person visits and accelerated the adoption of remote monitoring solutions.^2^ Studies such as the HOME and MARINA studies have demonstrated that early detection of choroidal neovascularization (CNV) in AMD is associated with better visual acuity outcomes, underscoring the need for innovative monitoring strategies.^2,28^ Nevertheless, long-term adoption of any one specific solution has been limited. Previous research has validated the utility of preferential hyperacuity perimetry (PHP) devices in detecting CNV conversion earlier than standard care, with patients retaining better visual acuity at diagnosis.^2^ However, these studies often involved controlled cohorts with limited real-world applicability. The Macustat test is a multi-modal scan of the central retinal function which includes visual acuity testing, dynamic digital Amsler grid and linear hyperacuity test designed to provide a comprehensive interrogation of foveal vision for the early detection of pre-symptomatic defects such as scotomas and metamorphopsias.^20^ When integrated with a care management and patient engagement service it can be used to enhance the standard-of-care episodic clinic paradigm in the management of chronic eye diseases through early detection and timely intervention. This multimodal design offers flexibility for daily, weekly, or monthly self-testing and, when coupled with structured PCM-enabled vision-coach support, directly addresses the adherence and retention barriers that have limited prior remote monitoring tools.

This is the first real-world evidence-based population health study of clinical care outcomes from a novel remote physiologic monitoring and principal care management (RPM-PCM) program for patients with chronic retinal conditions. By integrating validated digital diagnostics with proactive care-management infrastructure, the approach demonstrates how remote monitoring can be translated into sustainable, scalable programs that ensure care continuity and optimize visual outcomes.

## Methodology

This single-arm real-world population health intervention study evaluated RPM and PCM outcomes using the RemoniHealth platform Macustat over 18 months (January 1, 2024, to June 1, 2025) across 33 community-based retinal specialist practices in the United States, involving 2,216 patients and 82,644 encounters. We previously demonstrated the validity of the Macustat test here: https://journals.sagepub.com/doi/10.1177/20552076221132105.

Eligible patients had sight-threatening chronic retinal conditions (e.g., AMD, DR, RVO) likely to progress between follow-ups, were enrolled in the study. Patients were diagnosed through a clinical fundoscopic exam by a retina specialist with confirmatory imaging via spectral-domain optical coherence tomography (SD-OCT) or fundus fluorescein angiography. Inclusion criteria included age ≥18 years, access to the RemoniHealth platform, and informed consent. Exclusions comprised of unstable systemic conditions and lack of confirmed diagnosis. Of 2,824 screened patients, 2,216 (78.5%) were enrolled, with 608 excluded due to non-qualifying conditions or consent issues.

The RPM-PCM service provided 24/7 access to a HIPAA-compliant digital platform featuring the Macustat test, assessing visual acuity (tumbling E optotypes), contrast sensitivity, and metamorphopsia (dot alignment tasks) via personal devices such as smart phones and tablets. Recommended testing frequency was 2-3 times weekly. RPM monitored metamorphosia changes, visual fields, and symptoms, generating alerts for deviations >2 SD from baseline. PCM involved a triage team (certified nurses, specialists) reviewing alerts, categorized as care management (e.g., symptom reports) or Macustat® flags (e.g., metamorphopsia changes), with secure communication to facilitate interventions. Progression alerts are defined as a reading change in the multi-modal retinal function scan.

Data from the RPM-PCM database included demographics, diagnoses, encounters, alerts, and interventions. In-office visual acuity (Snellen, converted to logMAR) and SD-OCT data collected after a test was flagged were used to validate Macustat results. Adherence was calculated as the percentage of completed sessions, with an average compliance rate of 72.1%. Patient months of active monitoring totaled 16,805, with 27,897 total months.

Descriptive statistics summarized data: continuous variables (e.g., age, tests/patient) as means (SD), categorical variables (e.g., diagnosis) as percentages. The true positive rate from confirmed flags was calculated with 95% CI (Clopper-Pearson). Subgroup analyses by diagnosis and adherence used chi-square tests (p<0.05). R software (v4.2.1) and Python (v3.10) ensured data integrity via double-entry verification. The study adhered to the Declaration of Helsinki, and Institutional Review Board (IRB)/Ethics Committee approval was obtained (Elemental IRB #CAN-012). The study was listed on clinicaltrials.gov: NCT07216677.

## Results

A total of 82,644 encounters were recorded for 2,216 patients (mean age 77.9 years, SD 8.43; 65.9% female) over 18 months, averaging 33.82 tests/patient (SD 36.9) (Table 1). Diagnoses included 47.1% AMD (1,248 patients; 71.4% dry, 28.6% CNV), 43.6% DR, 25.6% RVO, 17.2% epiretinal membrane, and others (e.g., 2.9% myopic CNV) (Table 2). Patient months of active monitoring were 16,805, with 27,897 total months.

**Table 1.** Study timeline and Patient Demographics.

|  | N or % | Notes |
| --- | --- | --- |
| Period | 01/01/2024-06/01/2025 |  |
| Period (months) | 18 |  |
| Total Sites/ Doctors | 33 |  |
| Age (mean) | 77.9 | SD 8.43 |
| Gender Female | 65.90% |  |
| Total Monitoring Encounters for period | 82,644 |  |
| Total Monitored Patients for the period | 2,216 |  |
| Total Patient Months of Active Monitoring | 16,805 |  |
| Total Patient Months of Monitoring | 27,897 |  |
| Mean number of tests per patient | 33.82 | SD 36.9 |
| Mean number of tests per patient in 18-month period | 33.82 | SD 36.9 |
| Patients with at least one test every month (18/18) | 179 | 8.26% |
| Average compliance rate (engaged patients) | 72.10% | SD 30.5% |
| Compliance rate 100% | 47.8% |  |
| Compliance rate 80% or higher | 60.2% |  |

**Table 2.** Patient Disease Demographics.

| *Disease Category* | *Percentage (%)* | *Notes* |
| --- | --- | --- |
| Age-Related Macular Degeneration (AMD) (1,248) | 47.10% | *Dry AMD 71.4% and CNV 28.6%* |
| Diabetic Retinopathy | 43.60% |  |
| Retinal Vein Occlusion (RVO) | 25.60% |  |
| Epiretinal Membrane / Macular Pucker | 17.20% |  |
| Central Serous Chorioretinopathy | 1.70% |  |
| Myopic CNV | 2.90% |  |
| Myopic Degeneration | 1.30% |  |
| Monitoring for Macular Toxicity | 1.70% |  |
| Other Macular / Posterior Pole Diseases | 8.80% |  |
| Miscellaneous (e.g., Myopic CNV, CSCR) | 5.90% |  |
| Cataract | 3.40% |  |
| Glaucoma | 2.90% |  |

Mean compliance was 72.1% (SD 30.5%), with 44.1% (955/2,216) achieving ≥80% adherence and 8.26% (179/2,216) testing monthly (Table 1). High-adherence patients (>80%) showed a 2.1-fold higher true positive rate (OR 2.1, 95% CI 1.3-3.4, p=0.002). Preliminary visual acuity stabilized in 68% of treated patients (mean change -0.02 logMAR), with 22% improving (+0.15 logMAR) and 10% worsening. VFQ-25 scores improved by 7.2 points (SD 5.1, p<0.001) at 6 months.

The platform generated 241 alerts, with 56.2% (135) were care management, non-test related alerts, 41.9% (101) Macustat® test flags, and 2.1% (5) missing/other (Table 3). Of 101 flags, 73 were resolved (56 true positives, 77% true positive rate, 95% CI 66-86%; 17 false positives, 23%), with 22 unknown. The annualized detection rate was 4 detections per 100 patient-years. True positive flags led to interventions in 56 patients (2.5% of cohort), with 93% (52/56) receiving intravitreal (IVT) injections (65% aflibercept, 25% ranibizumab, 10% bevacizumab). AMD accounted for 52% of flags (52/101), DR 38% (38/101), and RVO 10% (11/101).

**Table 3.** Macustat Alert Characteristics and Outcomes.

|  | N or % | Notes |
| --- | --- | --- |
| Total Alerts | 241 |  |
| Total Patients receiving an alert | 205 |  |
| Care Management Alerts (Courtesy) | 135 | 56.20% |
| Macustat ® Monitoring Flags | 101 | 41.90% |
| Missing/ Other | 5 | 2.10% |
| True Positive Flags | 56 | 77% |
| False Positive Flags | 17 | 23% |
| Unknown | 22 |  |
| Annualized Detection Rate (per patient-year of active testing) | 4 | *detections per 100 patient years* |
| Rate of IVT injections for all true positive flags | 93% |  |

To improve patient adherence and testing fluency, the RemoniHealth Care management team employs vision coaches and ophthalmic professionals to address questions related to Macustat test taking. Of all patients included in this study, 76% had test related questions, clinic scheduling and testing follow-up questions, and 21% had access to care questions. 49% of the patients sought help related to ocular symptoms while taking the Macustat, and 58% asked for disease education upon follow-up with testing (Table 4).

**Table 4.** Frequency of RPM/Care Management Consults.

| Care Management/PCM Issues | *% of pts* |
| --- | --- |
| Test related questions | 76.0% |
| Patient symptoms (ocular discomfort, dry eye, etc..) | 49.0% |
| Disease education | 58.0% |
| Clinic scheduling and follow-up question | 32.0% |
| Access to care questions | 21.0% |
| Other | 38.0% |

In a sub-study survey of 34 subjects using the RPM service with at least 60 days of remote monitoring with the Macustat visual function test, patients reported high levels of usability, confidence, and overall satisfaction. The vast majority (88%) indicated that the test was easy to perform, with more than three-quarters (76%) regarding participation as worthwhile and nearly 60% considering a frequency of one to two sessions per week to be adequate. Importantly, 65% of patients reported improved peace of mind with remote monitoring, 53% expressed increased confidence in their treating physician, and 59% believed that integration of remote testing provided superior care compared to in-office visits alone. Perceived impact on the doctor–patient relationship was also favorable, with 47% of patients feeling more connected to their physician and 35% suggesting that availability of such a program would influence their choice of provider. Overall evaluations of the platform were positive: 76% assigned a rating of ≥4 out of 5 (mean score 3.94), while 71% expressed a high likelihood of recommending the program to others (≥8 out of 10; mean score 7.71). 29% rated the program as equally important as in-office consultations.

## Discussion

This single-arm real-world population health intervention study, encompassing 82,644 monitored encounters across 2,216 patients, robustly validates the efficacy of the RemoniHealth RPM-PCM platform in enhancing the remote monitoring and management of chronic retinal diseases. The integrated digital health monitoring system, coupled with the principal care management service, is designed to extend the conventional clinic-based retina patient management for care continuity. The results demonstrate effective real-world patient usability, adoption and engagement with long-term adherence—particularly noteworthy in a predominantly elderly cohort (mean age 77.9 years). Conducted in a community, non-study setting, the platform exhibits high sensitivity for detecting asymptomatic disease progression, augmenting the clinical outcomes and utility associated with the current standard-of-care with episodic office visits. Indeed, out of the 101 patients that were successfully flagged by the Macustat, only 12 reported visual symptomology (e.g. awareness of metamorphopsia, scotoma or decreased visual acuity) at the time of alert, resulting in early treatment interventions that would have otherwise been delayed. For conditions such as AMD and diabetic retinopathy, which are leading causes of irreversible blindness and where timely interventions (e.g., anti-VEGF therapy) have been shown to minimize visual impairment, the finding that RPM-PCM enhances disease management and patient care with timely detection and interventions in 93% of incident cases underscores its potential significance from population health perspective.

A distinctive feature of this investigation is its novel patient engagement and care continuity RPM-PCM model, uniquely combining digital health diagnostic and monitoring technology with proactive, high-touch outreach from vision coaches. This synergistic approach fosters robust adherence and engagement, yielding a 77% true positive rate for detecting asymptomatic progression and an annualized detection rate of 4 per 100 patient-years, a performance that compares favorably with prior home-monitoring tools. For example, in the randomized HOME trial of preferential hyperacuity perimetry (PHP), earlier CNV detection preserved more vision at conversion vs standard care,^2^ whereas subsequent real-world analyses of ForeseeHome showed mixed positive predictive performance and workflow challenges.^21^ Large UK diagnostic-accuracy work (MONARCH) similarly found that several home vision tests had limited diagnostic accuracy for detecting active nAMD in routine follow-up.^11,18^ Mobile-app approaches (e.g., OdySight, Alleye) demonstrate feasibility but variable retention and heterogeneous accuracy across studies.^13,22,15^ In parallel, home OCT has recently advanced from feasibility to FDA De Novo authorization (Notal Vision SCANLY, 2024) with pivotal trials showing patient-operated imaging and AI-assisted grading that align with in-clinic OCT for AMD monitoring,^23^ FDA clinical trial ID: NCT04907409, NCT05202587]. The diverse, non-homogeneous patient population, encompassing a broad spectrum of retinal conditions (47.1% AMD, 43.6% diabetic retinopathy, 25.6% RVO), aligns with clinical performance benchmarks from controlled trials, such as the HOME study’s preferential hyperacuity perimetry (PHP) for AMD and the ETDRS, which reported 90% vision preservation with early intervention. The 93% IVT injection rate among true positives further highlights the platform’s ability to translate early detection into actionable clinical interventions.

The multi-modal Macustat test, previously validated for remote assessment of central retinal function,^20^ leverages visual acuity, digital Amsler grid, and linear hyperacuity assessments, enabling remote self-administration virtually on consumer personal devices at the patient’s convenience. This user-friendly design, tailored for an elderly population (mean age 77 years), achieved an average of 33.82 tests per patient over 18 months, reflecting high usability and engagement, which compares favorably with engagement reported for other vision-function apps such as OdySight and Alleye, where real-world retention and performance have been variable across studies.^22,13,24^ Notably, while classic preferential hyperacuity perimetry (PHP) in the randomized HOME trial demonstrated earlier CNV detection with preserved vision versus standard care,^2^ subsequent real-world and diagnostic-accuracy analyses highlight mixed PPV and false-alarm burdens for home hyperacuity tools and limited accuracy for several app-based tests, underscoring the need for robust engagement plus confirmatory pathways.^11,18^ Within this context, Macustat’s consumer-device, multimodal design (acuity + Amsler + hyperacuity) offers practical flexibility for daily/weekly/monthly self-testing—complementary to emerging home OCT systems that provide anatomical fluid visualization but require dedicated hardware,^23^ FDA clinical trial ID: NCT04907409, NCT05202587. Digital Amsler implementations also continue to mature, with recent validations supporting their role in remote macular screening/monitoring when paired with structured follow-up.^11,18^

Our study has a 77% true positive rate which compares favorably with prior remote monitoring technologies. ForeseeHome, with an 85% sensitivity in a controlled AMD cohort, underperformed in real-world settings (6.8% true positive rate), as noted in the 2021 Ophthalmology Retina study, due to limited applicability and high false positives.^2^ MONARCH’s digital monitoring achieved 74% sensitivity for nAMD but was constrained to a narrow population with 26% uptake barriers.^11^ Odysight exhibited low retention (43% at 3 months, 24% at 9 months), reflecting adherence challenges, while Alleye and My Vision Track, though promising with hyperacuity testing, lack the scale and multi-disease scope of RemoniHealth.^13,15^ Taken together, these comparisons emphasize the value and utility of this combined digital health + care-management model with dedicated vision-coach outreach, which appears to overcome common barriers of sensitivity, adherence, and scalability, thereby supporting more effective real-world disease monitoring.

Despite its strengths, this study, among the first and largest investigations of remote physiologic monitoring in combination with principal care management, has several limitations inherent to its real-world design. The lack of specificity in patient selection and enrollment, the non-protocolized disease management across the different eye care providers and the lack of feedback from the electronic health record to the clinical data for all the patients, limits our ability to analyze the performance metrics more comprehensively and explore subgroup analyses. This is particularly true for our ability to confirm all incident cases, and in particular, the cases that were not detected. We do not have the data to determine false negative rate (lack of information on patients without flags due to the real-world nature of the study) although we know that whatever the rate is, it is the standard-of-care today associated with unmonitored episodic clinic visits without remote care RPM and PCM. Further, a quality-of-life instrument could have been useful in complementing the patient satisfaction results to gain deeper insights. Additional data such as in-clinic BCVA after an alert, time between alert and the next scheduled retina visit, data on patients without alerts over the course of the study would have strengthened our findings. These gaps can be addressed in future investigations and with more robust data access.

In conclusion, this study validates a novel RPM-PCM care continuity system as an extension to the eye care clinics for the remote management of chronic retinal diseases. The system could improve care outcomes with early detection of asymptomatic progression and timely intervention while delivering robust patient engagement and adherence.

**Figure 1.**
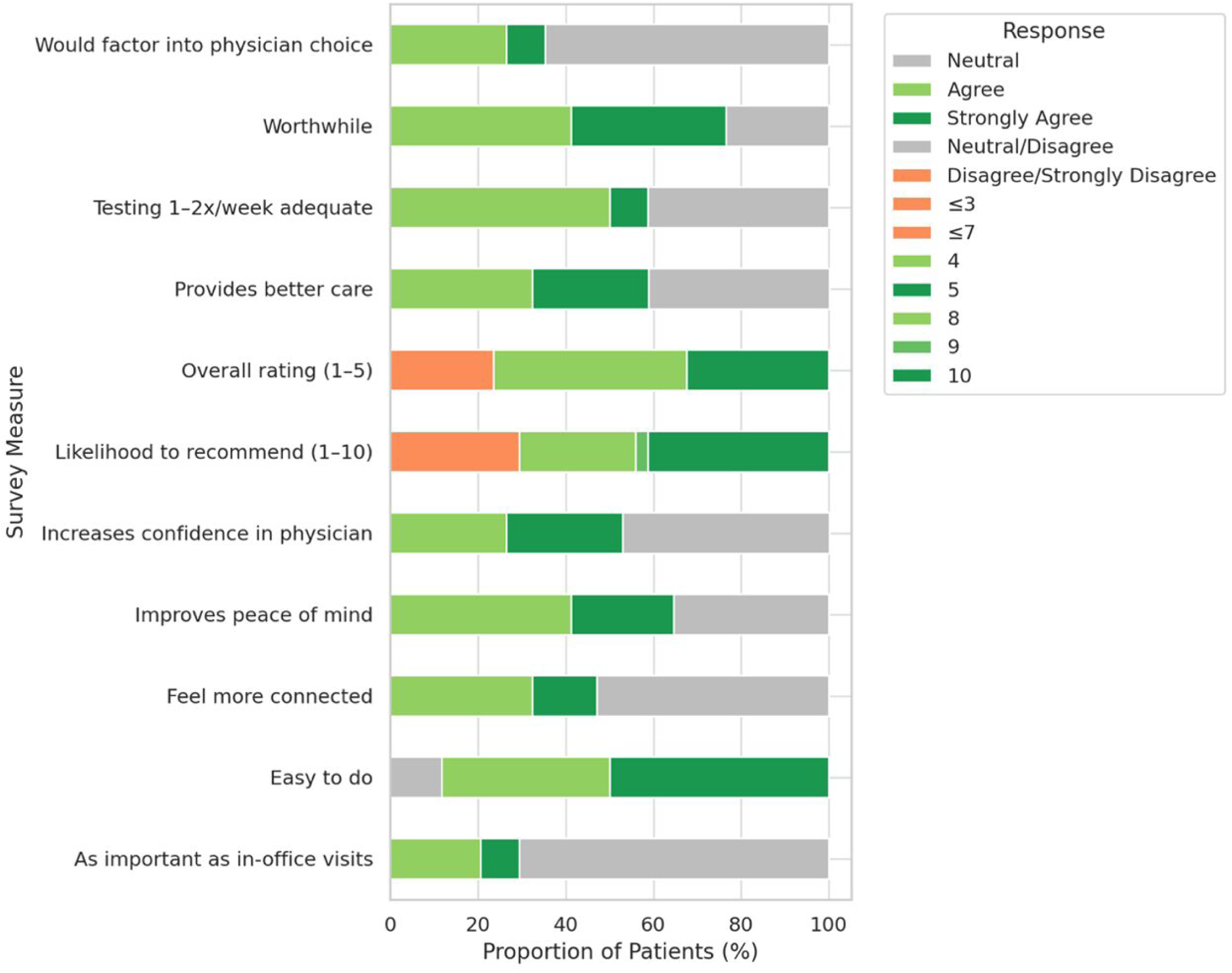
Patient Satisfaction Survey Results (N = 34).

## Data Availability

All data produced in the present study are available upon reasonable request to the authors.

